# Comparing Developmental Outcomes of Autistic Preschoolers Across Special and Mainstream Educational Settings

**DOI:** 10.64898/2026.08.23.26361140

**Authors:** Moran Bachrach, Michal Ilan, Michal Faroy, Analya Michaelovsky, Dikla Zagdon, Yair Sadaka, Omer Bar Yosef, Adi Aran, Michal Begin, Ditza Zachor, Einat Avni, Judah Koller, Idan Menashe, Tamar Kolodny, Ilan Dinstein, Gal Meiri

## Abstract

In many high-income countries, autistic children attend preschools ranging from exclusive special education (SE) to inclusive mainstream education (ME). These settings differ in staff expertise, capacity to implement structured autism interventions, exposure to typically developing peers, and cost. In this prospective longitudinal study, we compared 119 autistic children across three preschool settings in southern Israel: SE with TABAM services, an extended intervention program; SE without TABAM; and ME. Children completed behavioral assessments at the beginning and end of their first preschool year, yielding measures of cognition, autism symptom severity, joint attention, verbal abilities, adaptive behaviors, and aberrant behaviors. Developmental trajectories varied across children, with some demonstrating marked gains and others showing limited progress. On average, developmental changes were modest across most domains and were not explained by educational setting. The only exception was verbal ability, where children in SE with TABAM showed greater gains than children in SE without TABAM. These findings suggest that autistic children in ME and SE demonstrated broadly similar developmental trajectories during their first preschool year. Further large-scale research is needed to identify which children may benefit more from specific educational environments and intervention approaches, and to inform ongoing efforts to optimize preschool services for autistic children.

## Introduction

Children diagnosed with autism are remarkably heterogeneous, exhibiting varying levels of social difficulties, restricted and repetitive behaviors (RRBs), cognitive abilities, language skills, adaptive behaviors, and aberrant behaviors (Mandelli et al., 2024; Trembath & Vivanti, 2014; Zachor & Ben-Itzchak, 2017). In most high-income countries, preschool children with autism, 2-5 years old, are placed in preschool settings that vary along a continuum from inclusive mainstream education (ME) to exclusive special education (SE). There are currently no guidelines for placing preschool children with specific abilities or difficulties in different settings (Rattaz et al., 2020; Towle et al., 2018; White et al., 2007) and there is considerable overlap in the autism severity, adaptive behaviors, cognitive, and language abilities of children across settings (Bachrach et al., 2026; Ilan et al., 2021).

In Israel, publicly funded SE and ME preschool settings differ substantially. SE offers an exclusive environment where a small number of autistic children (∼9 children) receive structured autism-specific interventions from a highly professional multidisciplinary educational team who tailor the intervention to the specific needs of each child (Arnold et al., 2021). Many studies have demonstrated that early autism interventions improve later outcomes (Gabbay-Dizdar et al., 2022; Hyman et al., 2020; Zwaigenbaum et al., 2015), thereby motivating large investment in specialized preschool SE settings.

In contrast, inclusive ME preschools have larger classes (∼30 children) that are managed by a smaller educational team who are usually not trained to provide autism-specific intervention. Autistic children who are integrated in these settings often qualify for a personal aid to assist with daily activities and peer interactions (Barton et al., 2012; Lynch & Irvine, 2009), but teachers often report that integration is challenging (Gavaldá & Qinyi, 2012; Lindsay et al., 2014). Despite these challenges, autistic children in integrated settings benefit from exposure to their typically developing peers (Arnold et al., 2021b; Farrell, 2000; Sansour & Bernhard, 2018a). Finally, it is important to consider that SE settings are considerably more expensive to operate with the cost per child estimated as 4 times higher than ME (Chasson et al., 2007).

Studies examining the development of autistic children separately in each setting have reported that children improve in both. For example, children with autism placed in SE exhibited significant improvement in cognitive and social abilities, as well as in adaptive skills (Harris et al., 1991; Talbott et al., 2016; Zachor & Ben Itzchak, 2010). Similarly, children with autism placed in ME settings showed significant improvements in adaptive behaviors, communication abilities, language, and academic skills (Stahmer & Ingersoll, 2004;Little, 2017).

Remarkably, only a handful of studies have directly compared the development of autistic children across educational settings and have reported mixed results. One study of 97 autistic children, 4-16 years old, reported that children in SE and ME settings improved similarly in adaptive behaviors while children in SE made greater improvements in conduct and social abilities (Reed et al., 2012). Another study of 108 autistic children, 5-18 years old, reported that children in SE developed better language abilities than those in ME (Waddington & Reed, 2017). In contrast, a third study of 98 preschool autistic children, 3-5 years-old, reported that children in ME made larger cognitive gains than those in SE (Nahmias et al., 2014). Finally, a recent randomized controlled trial (RCT) reported that 44 preschool autistic children, 15–32 months old, exhibited similar cognitive, language, and social outcomes when placed in either SE or ME settings that implemented the Group Early Start Denver Model intervention with the aid of academic researchers (Vivanti et al., 2019). Note that the two later studies were the only ones to specifically study young autistic children in preschool settings, which is of particular interest given that early development is characterized by large neural plasticity that may enable more effective intervention (Nelson et al., 2024; Sullivan et al., 2014).

Hence, the objective of our study was to directly compare developmental trajectories of autistic children attending three publicly available preschool settings in Israel. In this prospective longitudinal study, we assessed cognitive abilities, expressive language abilities, autism symptom severity, joint attention (JA) skills, adaptive behaviors, and aberrant behaviors at the beginning and end of the children’s first year in preschool. We then compared longitudinal changes across SE and ME settings and examined whether developmental gains were associated with children’s initial abilities or difficulties.

## Methods

### Participants and procedures

We recruited 119 autistic children, 93 boys, mean age of 39.05 months (SD = 4.74), at the beginning of their first year in preschool. All recruited children were born between 2018 and 2020 and started their first year of preschool between 2021 and 2023, respectively. Participants were recruited through the Azrieli National Centre for Autism and Neurodevelopment Research (ANCAN), a collaborative effort between Ben-Gurion University of the Negev (BGU) and eight clinical sites where autistic children are diagnosed across Israel (Dinstein et al., 2020). Of these children, 23 (19.3%) were placed in inclusive ME, 55 (46.2%) were placed in exclusive SE settings with extended intervention (TABAM), and 41 (34.5%) were placed in exclusive SE without TABAM. All participating children were diagnosed with autism using DSM-5 criteria as determined by both a licensed developmental psychologist and a pediatric neurologist or child psychiatrist. All children were born without major complications at 36-42 weeks gestational age with birth weight >2500 grams, and no known metabolic, neurological, or genetic syndromes.

Children completed the following assessments at ANCAN within four months of their entry into preschool and again approximately one year later (Table 1). Autism Diagnostic Observation Schedule, 2^nd^ edition (ADOS-2) assessments were completed by 100 (84%) children at T1 and 89 (75%) at T2. Cognitive assessments were completed by 81 (68%) children at T1 and 73 (61.4%) at T2. Adaptive Behavior Rating Scale, 3^rd^ edition (ABAS-3) questionnaires were completed by parents of 73 (61.4%) children at T1 and 71 (60%) children at T2. Aberrant Behaviors Checklist (ABC) questionnaires were completed by parents of 67 (56.3%) children at T1 and 70 (59%) children at T2. The Soroka University Medical Center (SUMC) Helsinki Committee approved this study, and parents of all children signed informed consent.

**Table 1:** Characteristics of participating children in each educational setting.

|  | ME<br>(n=23) | SE<br>(n=41) | SE with Tabam<br>(n=55) | Overall<br>(n=119) |
| --- | --- | --- | --- | --- |
| Measures | Mean (SD) |  |  |  |
| Age at diagnosis (Months) | 33.8 (7.48) | 28.0 (12.0) | 29.5 (6.54) | 29.8 (9.13) |
| Age at T1 (Months) | 40.5 (4.25) | 39.1 (4.50) | 38.7 (5.09) | 39.2 (4.75) |
| Age at T2 (Months) | 52.0 (4.57) | 51.3 (4.79) | 51.6 (3.84) | 51.6 (4.31) |
| Sex, n (%) boys | 17 (73.9) | 32 (78) | 44 (80) | 93 (78.2) |
| Paternal education (Years) | 14.2 (2.61) | 13.3 (2.00) | 13.2 (2.44) | 13.4 (2.33) |
| Maternal education (Years) | 15.4 (2.58) | 13.4 (2.27) | 13.5 (2.51) | 13.9 (2.54) |
| Income | 1.85 (1.18) | 1.34 (0.90) | 1.19 (1.04) | 1.38 (1.05) |
| ADOS-2 total CSS at T1 | 6.15 (2.35) | 7.06 (2.31) | 7.23 (1.87) | 6.96 (2.13) |
| ADOS-2 total CSS at T2 | 5.71 (2.03) | 6.47 (2.27) | 6.82 (2.02) | 6.44 (2.13) |
| ADOS-2 SA-CSS at T1 | 5.85 (2.48) | 6.45 (2.49) | 6.50 (2.18) | 6.35 (2.33) |
| ADOS-2 SA-CSS at T2 | 5.10 (2.02) | 5.70 (2.48) | 6.21 (2.26) | 5.78 (2.30) |
| ADOS-2 RRB-CSS at T1 | 7.25 (1.68) | 8.39 (1.45) | 8.33 (1.42) | 8.13 (1.54) |
| ADOS-2 RRB-CSS at T2 | 7.52 (1.78) | 7.97 (1.43) | 8.16 (1.28) | 7.94 (1.46) |
| ADOS-2 verbal ability at T1 | 3.70 (1.84) | 5.23 (1.67) | 5.00 (1.52) | 4.81 (1.71) |
| ADOS-2 verbal ability at T2 | 2.43 (1.40) | 4.63 (1.88) | 3.74 (1.72) | 3.73 (1.88) |
| ADOS-2 JA score at T1 | 5.57 (4.13) | 8.16 (4.27) | 8.04 (3.53) | 7.56 (4.00) |
| ADOS-2 JA score at T2 | 4.47 (2.65) | 6.83 (3.81) | 6.62 (3.68) | 6.22 (3.61) |
| Cognitive score at T1 | 87.9 (24.9) | 59.9 (15.0) | 68.5 (20.6) | 69.9 (22.2) |
| Cognitive score at T2 | 93.3 (24.6) | 64.5 (20.6) | 66.3 (19.7) | 71.2 (23.6) |
| ABAS GAC score at T1 | 80.5 (16.3) | 60.7 (14.9) | 67.4 (17.4) | 68.1 (17.7) |
| Measures | Mean (SD) |  |  |  |
| ABAS GAC score at T2 | 77.8 (18.5) | 55.6 (14.8) | 64.3 (17.1) | 65.1 (18.5) |
| ABC Irritability score at T1 | 6.17 (7.40) | 18.3 (11.0) | 11.2 (10.1) | 13.1 (10.9) |
| ABC Irritability score at T2 | 9.05 (8.01) | 14.6 (11.5) | 12.8 (8.27) | 12.3 (9.40) |

ME: Mainstream Education. SE: Special Education. ADOS-2: Autism Diagnostic Observation Schedule. CSS: Calibrated Severity Scores. SA: Social Affect. RRB: Restricted and Repetitive Behaviors. JA: Joint Attention score. ABAS: Adaptive Behavior Rating Scale. ABC: Aberrant Behaviors Checklist. T1: First study time-point at entry into preschool. T2: Second study time-point at end of first year.

### Measures

#### ADOS-2

Children completed the toddler module or modules 1–2 of the ADOS-2 at T1 and modules 1-3 at T2 according to their age a language ability (Lord et al., 2012). The evaluation was conducted by an experienced clinician with research reliability. ADOS-2 raw scores were transformed into Calibrated Severity Scores (CSS), which allows comparison of autism severity across children of different ages, language capabilities, and over time (Esler et al., 2015; Gotham et al., 2009). Separate ADOS-2 CSS scores were computed for social affect (SA) and restricted and repetitive behaviors (RRBs) (Esler et al., 2015; Hus et al., 2014).

#### Cognitive assessments

Cognitive ability was measured using one of several cognitive tests as determined by a licensed developmental psychologist. The Bayley Scales of Infant and Toddler Development, Third Edition (Viezel et al., 2014a) was used with 15 (∼18.6%) children at T1 and 1 (∼1.3%) child at T2, the Mullen Scales of Early Learning (MSEL, Mullen, 1995) was used with 59 (∼72.8%) children at T1 and 69 (94.5%) children at T2, and the Wechsler Preschool and Primary Scale of Intelligence, Third Edition (Luiselli et al., 2013a) was used with 7 (∼8.6%) children at T1 and 3 (∼4.2%) children at T2. The three tests yield equivalent standardized scores with a mean of 100 and a standard deviation of 15. Since strong correlations exist between the Bayley and Wechsler tests, as well as between the Mullen and Bayley tests (Bayley, 2006; Lense et al., 2014a), we combined scores from these tests in our analysis.

#### Verbal abilities

Verbal ability was evaluated using the A1 item of the ADOS-2 assessment, which contains the clinician’s estimate of the child’s expressive language abilities. We used a previously developed 8-point scale to combine verbal ability estimates across ADOS-2 modules (Visser et al., 2017) with the following values:

0. Children use sentences in a largely correct fashion (complex utterances with >2 clauses).
1. Children exhibit relatively complex speech (occasional utterances with >2 clauses) with recurrent grammatical errors.
2. Children exhibit non-echoed speech with utterances of >3 words.
3. Children mainly use individual 2- to 3-word phrases, with or without minimal grammar.
4. Children mainly use individual words with occasional simple phrases.
5. Children only use individual words (minimum of 5 different words).
6. Children only use echoed speech (<5 words).
7. No language production at all.

#### Joint Attention (JA)

Following our previous work (Nitzan et al., 2026), we quantified JA by summing the scores of the following 6 items from the ADOS-2: Pointing (item A7 in the Toddlers module and Module 1, item A6 in Module 2), Gesturing (item A8 in the Toddlers module and Module 1 and item A7 in Module 2), Showing (item B12 in the Toddlers module, B9 in Module1, and B5 in Module 2), Initiating JA (item B13 in the Toddlers module, B10 in Module1, and B6 in Module 2), Unusual Eye contact (item B1 in all modules), and Reaction to JA (item B14 in the Toddlers module, B11 in Module1, and B7 in Module 2). In line with the ADOS algorithm, scores of 3 were changed to 2, and scores of 8 were changed to 0, yielding a total JA scale of 0-12 with higher scores indicating poorer JA abilities. JA scores were not extracted for children who completed module 3 (3 children at T2), because the items described above are not available there.

This JA scale was inspired by previous factor analytic studies demonstrating that the six selected items load onto a common ADOS factor (Gotham et al., 2007, 2008; Oosterling et al., 2010). According to ADOS-2 guidelines, a score of 0 was assigned only when the child demonstrated clear social-communicative intent, which is central to JA. For example, pointing received a score of 0 only when accompanied by eye contact to share attention toward a remote object. Thus, the selected items captured a composite of JA behaviors comparable to those assessed by established measures such as the Early Social Communication Scales (ESCS; Mundy et al., 2003) and the Communication and Symbolic Behavior Scales (CSBS; Wetherby et al., 2002), although without separating responsive and initiated JA components

#### Adaptive behaviors

Adaptive behaviors were measured using the ABAS-3 (Balboni et al., 2014; Kane & Oakland, 2015) which was completed by the parents. This questionnaire includes 241 items, which are rated on a scale from 0 (Is not able) to 3 (Almost /Almost Always). The questionnaire has 10 subscales that measure Communication, Community Use, Functional Pre-academics, Home Living, Health and Safety, Leisure, Self-Care, Self-Direction, Social, and Motor abilities. These are summed to create three composite scores: Conceptual (CON; Communication, Functional Academics, Self-Direction), Social (SOC; Leisure, Social), and Practical (PRAC; Self-Care, Home Living, Community Use, and Health and Safety) as well as a final overall General Adaptive Composite (GAC) score.

#### Aberrant behaviors

Aberrant behaviors were measured using the ABC (Aman, 1985) which was completed by the parents. The ABC includes 58 items using a four-point rating scale from zero (“not a problem”) to three (“severe in degree”). The questionnaire includes five subscales: (1) irritability; (2) social withdrawal; (3) stereotypical behavior; (4) hyperactivity; and (5) inappropriate speech.

#### Follow up questionnaire

Parents completed a follow up questionnaire that included questions about parent education levels, socioeconomic status, average household income, and the educational placement setting of their child during the study period.

### Data analysis and statistics

All statistical analyses were conducted using RStudio (RStudio Inc., Boston, MA). Children were divided into three groups based on their educational setting (ME, SE without TABAM, and SE with TABAM). We imputed missing data with a multiple imputation procedure using the Random Forests technique with 20 iterations as implemented in the *mice* package in RStudio (Van Buuren & Groothuis-Oudshoorn, 2011). To ensure that this imputation process did not introduce biases to the data, we compared all study variables between children with and without missing data. This analysis demonstrated that there were no significant differences (all ps > .05) across the two groups in any of the study variables (Supplementary Table S1). Hence, missing data were not systematically biased toward children with specific characteristics (e.g., low cognitive abilities).

To examine developmental trajectories across educational settings, longitudinal change scores were calculated for all outcome measures as the difference between Time 2 and Time 1 (T2–T1). In addition, to assess overall longitudinal change across the entire sample, separate analyses of covariance (ANCOVA) were conducted for each outcome measure, in which change scores (T2–T1) were entered as the dependent variables while controlling for age of diagnosis and maternal education. Accordingly, positive change values indicate improvement for ability measures (e.g., cognitive and adaptive behavior scores) and negative change values indicate improvement for difficulty measures (e.g., ADOS-2 and ABC scores).

One-way analyses of covariance (ANCOVA) were conducted to compare change scores across educational settings while controlling for age at autism diagnosis and maternal education as covariates. Post hoc group comparisons were performed using Tukey’s Honestly Significant Difference (HSD) test, and effect sizes were estimated using η². Linear regression models were used to explain individual differences in cognitive and language change scores. Predictors included educational setting, baseline (T1) measures, baseline ADOS-2 SA and RRB scores, age at autism diagnosis, maternal education, and interaction between educational setting and baseline ability. Goodness of fit was evaluated using the coefficient of determination (R²) and adjusted R². Statistical significance was set at α = .05 for all analyses.

#### Preregistration Statement

This study was not preregistered

## Results

First, we examined the development of the entire sample and did not find any significant longitudinal changes between T1 and T2, indicating that, on average, children did not improve or deteriorate in any of the examined developmental domains during their first preschool year. We used ANCOVA analyses to assess the significance of longitudinal changes while controlling for age at autism diagnosis and years of maternal education. There were no significant changes (T2–T1) in any of the ADOS-2 derived measures, including total CSS (b = −0.26, SE = 1.25, t = −0.21, p = .834, 95% CI [−2.73, 2.20]), SA CSS (b = 1.02, SE = 1.47, t = 0.70, p = .486, 95% CI [−1.88, 3.93]), and RRB CSS (b = −0.55, SE = 1.02, t = −0.54, p = .594, 95% CI [−2.58, 1.48]). Similarly, no significant changes were observed in JA skills (b = −1.95, SE = 1.80, t = −1.08, p = .281, 95% CI [−5.51, 1.62]) or verbal abilities (b = −0.64, SE = 0.75, t = −0.85, p = .396, 95% CI [−2.12, 0.84]).

There were also no significant changes in cognitive scores (b = 9.88, SE = 11.40, t = 0.87, p = .388, 95% CI [−12.70, 32.46]) or adaptive behavior, including ABAS GAC scores (b = 0.84, SE = 7.80, t = 0.11, p = .915, 95% CI [−14.61, 16.29]), ABAS Conceptual (b = −8.47, SE = 7.48, t = −1.13, p = .260, 95% CI [−23.28, 6.34]), ABAS Social (b = −17.80, SE = 9.82, t = −1.81, p = .072, 95% CI [−37.24, 1.64]), and ABAS Practical (b = −9.18, SE = 9.45, t = −0.97, p = .333, 95% CI [−27.90, 9.54]). Similarly, no significant changes were observed in any of the ABC subscale scores, including irritability (b = −7.42, SE = 6.40, t = −1.16, p = .249, 95% CI [−20.10, 5.27]), social withdrawal (b = 2.70, SE = 5.32, t = 0.51, p = .612, 95% CI [−7.83, 13.24]), stereotypy (b = −1.27, SE = 2.73, t = −0.47, p = .641, 95% CI [−6.67, 4.13]), hyperactivity (b = −8.34, SE = 6.66, t = −1.25, p = .213, 95% CI [−21.53, 4.85]), and inappropriate speech (b = −3.07, SE = 2.16, t = −1.43, p = .157, 95% CI [−7.35, 1.20]). Age at autism diagnosis and years of maternal education were not significantly associated with change in any of the measures (all ps > .05).

### Longitudinal changes across educational settings

While there were no significant longitudinal changes, on average, there was large variability across autistic children with some exhibiting dramatic improvements while others exhibited considerable deterioration (Figure 1). To determine whether differences across children were associated with their placement in a particular education setting, we performed similar ANCOVA analyses while comparing changes across the three settings. Changes in ADOS-2 scores (T2-T1) did not differ significantly across the three educational settings. This was true for ADOS-2 Total CSS (F(2, 114) = 0.20, p = .823), ADOS-2 SA CSS (F(2, 114) = 0.10, p = .904), ADOS-2 RRB CSS (F(2, 114) = 1.92, p = .152), or ADOS-2 JA scores (F(2, 114) = 0.06, p = .943). Similarly, there were no significant differences across groups when comparing longitudinal changes in cognitive scores (F(2, 114) = 0.05, p = .951), ABAS GAC scores (F(2, 114) = 0.36, p = .698), ABAS conceptual scores (F(2, 114) = 0.70, p = .497), ABAS social scores (F(2, 114) = 0.42, p = .661), or ABAS practical scores (F(2, 114) = 0.52, p = .597). Longitudinal changes in ABC scores also did not differ significantly across groups in the irritability (F(2, 114) = 2.09, p = .128), social withdrawal (F(2, 114) = 1.49, p = .231), stereotypical movements (F(2, 114) = 0.86, p = .427), hyperactivity/noncompliance (F(2, 114) = 0.45, p = .638), or inappropriate speech (F(2, 114) = 1.13, p = .326) subscales.

**Figure 1:**
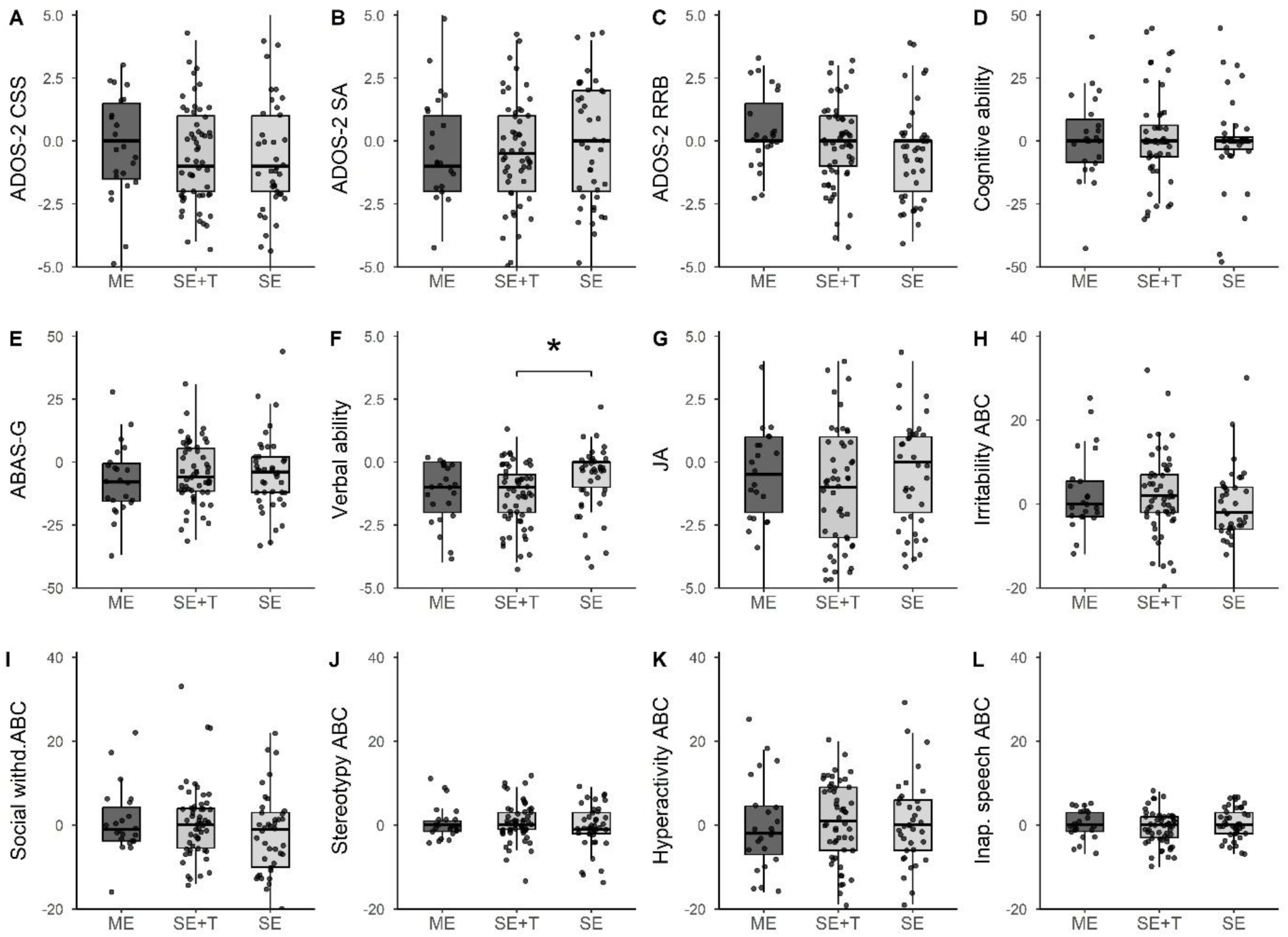
Comparison of longitudinal changes (T2-T1) across educational settings. Box and whisker plots present the distribution of longitudinal change in each variable for children in each educational setting. **A.** ADOS-2 CSS. **B.** ADOS-2 SA-CSS. **C.** ADOS-2 RRB-CSS. **D.** Cognitive scores. **E.** ABAS general scores. **F.** Verbal ability scores. **G.** JA scores. **H.** ABC irritability scores. **I.** ABC social withdrawal scores. **J.** ABC stereotypical behavior scores. **K.** ABC hyperactivity/noncompliance scores. **L.** ABC inappropriate speech scores. Asterisks: Significant difference across groups (* p<0.05, ** p<0.001). Note: Negative change scores indicate improvement for symptom measures (ADOS-2, JA, language, ABC), but for ABAS, positive change reflects improvement.

The only significant difference was found in verbal ability scores (F(2, 114) = 3.08, p = .050, η² = .05). Post-hoc Tukey comparisons revealed that children in special education with TABAM showed significantly greater improvement than those in special education without TABAM (mean difference = 0.65, 95% CI [0.02, 1.28], p = .041). No significant differences were observed between children in mainstream education and either special education group (TABAM vs. mainstream: mean difference = –0.16, 95% CI [–0.92, 0.59], p = .864; special without TABAM vs. mainstream: mean difference = 0.49, 95% CI [–0.31, 1.28], p = .317). Across all models, neither age at diagnosis nor maternal years of education was significantly associated with outcomes (all ps > .20).

### Longitudinal score changes

#### Predicting Cognitive and Verbal Ability Changes

Next, given the substantial developmental variability observed across children, we performed two multi-variate regression analyses to identify factors at T1 that were associated with greater gains in cognitive (Model A) or verbal (Model B) abilities. Using the first model we attempted to explain changes in cognitive scores with the following variables: educational setting (ME, SE, SE+T), cognitive score at T1, ADOS-2 SA-CSS at T1, ADOS-2 RRB-CSS at T1, age at diagnosis, year of maternal education, and the interaction between educational setting and cognitive score at T1 (Table 2). This model explained ∼25% of the variance in cognitive score changes and revealed that cognitive scores and ADOS-2 SA-CSS scores at T1 were the only significant predictors of cognitive improvement. Specifically, lower scores at T1 (i.e., poorer cognition along with higher social abilities) were associated with larger cognitive gains over time. Educational setting and the interaction between educational setting and cognition at T1 were not significant.

**Table 2:** Linear regression analysis explaining changes in cognitive abilities (Model A)

| Variable | b | SE | t | p | 95% CI |
| --- | --- | --- | --- | --- | --- |
| **Intercept** | 4.99 | 3.95 | 1.26 | .209 | [-2.75, 12.73] |
| Education setting: SE vs ME | -7.91 | 5.14 | -1.54 | .127 | [-17.98, 2.16] |
| Education setting: SE+T vs ME | -2.86 | 4.79 | -0.60 | .552 | [-12.25, 6.53] |
| **ADOS-2 SA-CSS at T1** | **-2.27** | **0.81** | **-2.79** | **.006\*\*** | **[-3.86, -0.68]** |
| ADOS-2 RRB-CSS at T1 | -0.79 | 1.22 | -0.65 | .518 | [-3.18, 1.60] |
| Age at diagnosis (M) | -0.08 | 0.19 | -0.43 | .668 | [-0.48, 0.30] |
| Maternal education (Y) | 0.61 | 0.83 | 0.74 | .464 | [-1.02, 2.25] |
| **Cognitive score at T1** | **-0.36** | **0.16** | **-2.28** | **.025\*** | **[-0.68, -0.05]** |
| SE × Cognitive score at T1 | -0.23 | 0.23 | -0.98 | .329 | [-0.69, 0.23] |
| SE+T × Cognitive score at T1 | -0.16 | 0.22 | -0.74 | .463 | [-0.60, 0.27] |

Observations: 119
$R^2 = 0.253$ ; Adjusted $R^2 = 0.192$
RMSE = 17.78
\* $p < 0.05$ .

Using the second model we attempted to explain changes in verbal ability with the same variables described above while replacing cognitive scores with verbal ability scores (Table 3). This model explained ∼23% of the variance in verbal ability changes and revealed that verbal ability and ADOS-2 SA-CSS scores at T1 were the only significant predictors of verbal ability changes. Specifically, lower verbal and ADOS-2 SA-CSS scores at T1 (i.e., better verbal and social function at T1) were associated with larger verbal ability gains over time (i.e., reduction in verbal scores). Educational setting and the interaction between educational setting and verbal abilities at T1 were not significant.

**Table 3:** Linear regression analysis explaining changes in verbal ability (Model B)

| Variable | b | SE | t | p | 95% CI |
| --- | --- | --- | --- | --- | --- |
| **Intercept** | **-1.53** | **0.38** | **-4.06** | **.001\*\*** | **[-2.27, -0.79]** |
| Education setting: SE vs ME | 0.82 | 0.43 | 1.90 | .060 | [-0.02, 1.67] |
| Education setting: SE+T vs ME | 0.15 | 0.41 | 0.38 | .707 | [-0.65, 0.96] |
| **ADOS-2 SA-CSS at T1** | **0.14** | **0.05** | **2.55** | **.012\*** | **[0.03, 0.25]** |
| ADOS-2 RRB-CSS at T1 | 0.09 | 0.09 | 0.93 | .356 | [-0.10, 0.27] |
| Age at diagnosis (M) | -0.02 | 0.01 | -1.80 | .075 | [-0.03, 0.00] |
| Maternal education (Y) | -0.03 | 0.05 | -0.73 | .465 | [-0.13, 0.06] |
| **Language abilities at T1** | **-0.50** | **0.19** | **-2.60** | **0.01\*** | **[-0.88, -0.12]** |
| SE × Language abilities at T1 | 0.28 | 0.21 | 1.34 | .183 | [-0.14, 0.70] |
| SE+T × Language abilities at T1 | 0.16 | 0.21 | 0.77 | .443 | [-0.25, 0.57] |

Observations: 119
$R^2 = 0.23$ ; Adjusted $R^2 = 0.17$
RMSE = 1.19
\* $p < 0.05$ .

## Discussion

This study aimed to compare developmental trajectories of young autistic children attending three publicly funded preschool settings in Israel during their first preschool year. The most notable finding was observed in the language domain, where children attending SE settings that included TABAM services demonstrated greater gains in verbal ability than children attending SE settings without TABAM. Beyond the language domain, developmental changes in cognitive abilities, adaptive functioning, autism symptom severity, JA, and aberrant behaviors were broadly similar across educational settings (Figure 1). These findings reveal the considerable heterogeneity characterizing developmental trajectories in autism and suggest that factors beyond educational placement contribute substantially to individual outcomes. Multivariate regression analyses revealed that initial cognitive, social, and language ability scores explained approximately one-quarter of the variability in cognitive and language developmental changes (Tables 2 and 3), regardless of the child’s educational setting. Overall, these findings suggest that individual differences play a meaningful role in shaping developmental trajectories and support the need for a more personalized approach to educational and intervention planning.

Among all developmental domains assessed, language ability stood out as the only area with significant differences across educational settings. On the one hand, the developmental trajectories observed in the inclusive cohort support the view that environments where preschool autistic children have opportunities to interact socially with typically developing peers can support language development (Fasano et al., 2023). This is the main motivation behind peer-mediated interventions and naturalistic developmental models, which suggest that rich, everyday exposure to socially engaged peers can scaffold communicative growth and support emerging expressive language abilities (Odom et al., 2021; Wong et al., 2015). On the other hand, the notably greater language gains observed in children placed in SE with TABAM aligned with prior literature suggesting that structured, intensive autism-specific interventions yield significant benefits for expressive language development. Crucially, TABAM settings often provide a more intensive framework for implementing Augmentative and Alternative Communication (AAC) techniques, which may yield even larger language benefits when integrated into high-dosage curricula (Pope et al., 2024; Sandbank et al., 2020). Furthermore, recent evidence suggests that while the relationship between intervention amount and overall outcomes is complex, increased intensity is often associated with more robust gains in specific communicative domains (Sandbank et al., 2024).

However, the broader pattern of findings revealed remarkably similar developmental trajectories across educational settings in most domains. These findings expand previous studies from our lab (Ilan et al., 2023) and others (Nahmias et al., 2014; Vivanti et al., 2019) reporting that children in inclusive ME preschools exhibit comparable developmental changes to children placed in exclusive SE preschools. Despite substantial differences in educational structure, intervention intensity, and available resources, children across settings demonstrated similar changes in cognitive abilities, adaptive functioning, autism symptom severity, joint attention, and aberrant behaviors. These findings suggest that no single educational setting appears uniformly superior across developmental domains during the first preschool year. Importantly, the similarity in outcomes across settings does not imply that developmental trajectories were uniform across children.

The large developmental heterogeneity apparent across children was explained to some degree (∼25%) by their initial abilities, as also reported by others (Brignell et al., 2018; Howlin et al, 2013; Vivanti et al., 2018, 2019). For example, higher initial social abilities were associated with greater gains in both cognitive and language domains (Mundy & Newell, 2007; Bottema-Beutel, 2016). Nevertheless, three-quarters of the variability in developmental trajectories across children was not explained by any of the variables examined in the current study. Thus, additional research is necessary to identify child, family, and intervention-related factors that shape early developmental trajectories and may influence responsiveness to different educational and therapeutic environments.

### Implications for Educational Placement Decisions

From a clinical and systems perspective, the current findings emphasize the importance of maintaining flexibility in educational placement decisions. Given the considerable variability observed across individual developmental trajectories, placement decisions should not rely solely on initial assessments or assumptions regarding the superiority of a particular setting. Rather, ongoing monitoring of developmental progress and periodic evaluation of intervention effectiveness may be essential for informing educational planning over time. The language gains observed in children receiving TABAM services further suggest that specific intervention components may be as important as the educational setting itself. Accordingly, future efforts may benefit from focusing not only on where children are placed, but also on which educational and therapeutic practices are implemented within those settings and for whom they are most effective.

### Context-Specific Policy and Limitations

These findings should be interpreted within the Israeli educational context, where there is a distinct separation between integrated ME and exclusive SE preschool settings. In other countries there are hybrid alternatives that combine partial integration in ME preschools with intensive early interventions in separate environments. In addition, approximately two thirds of autistic children in Israel are placed in SE preschools (Ilan et al., 2021; Ministry of health, 2014), while in other countries most autistic children are placed in inclusive ME preschools. Since this study was conducted in the south of Israel, which is a peripheral region with limited access to specialized services, our results may be limited to this geographic area.

Several additional limitations should be acknowledged. First, educational placement was not randomized and there were baseline differences across the three educational settings (Table 1). We, therefore, examined longitudinal changes rather than absolute outcomes and included baseline variables in our regression analyses to control for initial abilities. However, a randomized controlled trial would yield cleaner comparisons. Second, data collection occurred during both the COVID-19 pandemic and the October 7th war, which likely affected service availability and attendance. However, the extent to which these events differentially impacted educational settings could not be determined. Third, we measured developmental changes during a one-year period, which may not be long enough to reveal significant gains and intervention benefits that may require a longer period. Indeed, ADOS-2 calibrated severity scores may be less sensitive to short-term developmental changes (Hus et al., 2014; Gentles et al., 2024; Kim et al., 2018). Finally, the relatively small ME sample and lack of detailed intervention documentation should also be considered. Future studies with larger samples, longer follow-up periods, and precise measurements of intervention are needed to determine how preschools can be optimized to yield better outcomes for autistic children.

### Conclusions

This study provides empirical evidence from a community-based sample of preschool-aged autistic children in the south region of Israel, demonstrating marked individual developmental heterogeneity across ME and SE settings. The results suggest that children in integrated ME settings exhibit comparable developmental trajectories to those in exclusive SE settings, with the notable exception of language development, which is significantly enhanced by intensive intervention like TABAM. Large-scale studies aimed at identifying individual predictive factors and their interaction with specific educational settings are essential to further optimize resources devoted to autism preschool settings.

## Key Points

- Preschool autistic children in mainstream and special education settings exhibited substantial heterogeneity in developmental trajectories during their first preschool year.
- Developmental changes in cognition, adaptive behavior, autism symptom severity, joint attention, and aberrant behaviors were broadly similar across educational settings.
- Children attending special education settings that offered intensive autism-specific intervention demonstrated greater gains in verbal abilities than children attending standard special education settings.
- Initial cognitive, social, and language abilities explained a portion of the variability in developmental outcomes, regardless of educational setting.
- Future research is needed to identify child-specific characteristics associated with differential responses to educational and intervention environments.

## Data Availability

The data that support the findings of this study are not publicly available due to ethical and privacy restrictions but may be available from the corresponding author upon reasonable request and subject to institutional approval.

## Ethical Information

The study was approved by the Soroka University Medical Center Helsinki Committee [approval number: SOR-0222-14; approval date: 27 January 2015]. Parents or legal guardians of all participating children provided written informed consent prior to participation.

## References

Anderson, D. K., Lord, C., Risi, S., DiLavore, P. S., Shulman, C., Thurm, A., Welch, K., & Pickles, A. (2007). Patterns of growth in adaptive behavior in children with autism spectrum disorders. Journal of Abnormal Child Psychology, 35(2), 165–181. 10.1007/s10802-006-9066-y

Arnold, L., Milton, D., Beardon, L., & Chown, N. (2021). England and Autism. Encyclopedia of Autism Spectrum Disorders, 1774–1780. 10.1007/978-3-319-91280-6_102025

Bachrach, M., Ilan, M., Faroy, M., Michaelovsky, A., Zagdon, D., Sadaka, Y., Bar Yosef, O., Aran, A., Begin, M., Zachor, D., Avni, E., Koller, J., Menashe, I., Kolodny, T., Dinstein, I., & Meiri, G. (2026). Cognitive Abilities and Irritability Are the Main Factors Influencing Initial Placement of Autistic Preschoolers in Special or Mainstream Education in Israel. Autism Research, 19(3), e70188. 10.1002/aur.70188

Balboni, G., Tassé, M. J., Schalock, R. L., Borthwick-Duffy, S. A., Spreat, S., Thissen, D., Widaman, K. F., Zhang, D., & Navas, P. (2014). The Diagnostic Adaptive Behavior Scale: Evaluating its diagnostic sensitivity and specificity. Research in Developmental Disabilities, 35(11), 2884–2893. 10.1016/J.RIDD.2014.07.032

Barton, E. E., Lawrence, K., & Deurloo, F. (2012). Individualizing interventions for young children with autism in preschool. Journal of Autism and Developmental Disorders, 42(6), 1205–1217. 10.1007/s10803-011-1195-z

Bayley, N. (2006). Bayley scales of infant and toddler development (3rd ed.). San Antonio, TX: Harcourt Assessment.

Bottema-Beutel, K. (2016). Meta-analysis of joint attention associations with language outcomes in autism spectrum disorder. Journal of Autism and Developmental Disorders, 46(12), 3731–3741. 10.1007/s10803-016-2917-2

Brignell, A., Williams, K., Jachno, K., Prior, M., Reilly, S., & Morgan, A. T. (2018). Patterns and Predictors of Language Development from 4 to 7 Years in Verbal Children With and Without Autism Spectrum Disorder. Journal of Autism and Developmental Disorders, 48(10), 3282–3295. 10.1007/s10803-018-3565-2

Brinkley, J., Nations, L., Abramson, R. K., Hall, A., Wright, H. H., Gabriels, R., Gilbert, J. R., Pericak-Vance, M. A., & Cuccaro, M. L. (2007). Factor Analysis of the Aberrant Behavior Checklist in Individuals with Autism Spectrum Disorders. Journal of Autism and Developmental Disorders, 37, 1949–1959. 10.1007/s10803-006-0327-3

Chasson, G. S., Harris, G. E., & Neely, W. J. (2007). Cost Comparison of Early Intensive Behavioral Intervention and Special Education for Children with Autism. J Child Fam Stud, 16, 401–413. 10.1007/s10826-006-9094-1

Dinstein, I., Arazi, A., Golan, H. M., Koller, J., Elliott, E., Gozes, I., Shulman, C., Shifman, S., Raz, R., Davidovitch, N., Gev, T., Aran, A., Stolar, O., Ben-Itzchak, E., Snir, I. M., Israel-Yaacov, S., Bauminger-Zviely, N., Bonneh, Y. S., Gal, E., & Meiri, G. (2020). The National Autism Database of Israel: a resource for studying autism risk factors, biomarkers, outcome measures, and treatment efficacy. Journal of Molecular Neuroscience, 70(9), 1303–1312. 10.1007/S12031-020-01671-Z

Engelstad, A.-M., Holingue, C., & Landa, R. J. (2020). Early Achievements for Education Settings: An Embedded Teacher-Implemented Social Communication Intervention for Preschoolers With Autism Spectrum Disorder. Perspectives of the ASHA Special Interest Groups, 5(3), 582–601. 10.1044/2020_PERSP-19-00155

Esler, A. N., Bal, V. H., Guthrie, W., Wetherby, A., Weismer, S. E., & Lord, C. (2015). The Autism Diagnostic Observation Schedule, Toddler Module: Standardized Severity Scores. Journal of Autism and Developmental Disorders, 45(9), 2704–2720. 10.1007/s10803-015-2432-7

Farrell, P. (2000). The impact of research on developments in inclusive education. International Journal of Inclusive Education, 4(2), 153–162. 10.1080/136031100284867

Fasano, R. M., Mitsven, S. G., Custode, S. A., Debasish Sarker, |, Bulotsky-Shearer, R. J., Daniel, |, Messinger, S., & Perry, L. K. (2023). Automated measures of vocal interactions and engagement in inclusive preschool classrooms. 10.1002/aur.2980

Gabbay-Dizdar, N., Ilan, M., Meiri, G., Faroy, M., Michaelovski, A., Flusser, H., Menashe, I., Koller, J., Zachor, D. A., & Dinstein, I. (2022). Early diagnosis of autism in the community is associated with marked improvement in social symptoms within 1–2 years. Autism, 26(6), 1353–1363. 10.1177/13623613211049011/ASSET/IMAGES/LARGE/10.1177_13623613211049011-FIG4.JPEG

Gavaldá, J. M. S., & Qinyi, T. (2012). Improving the Process of Inclusive Education in Children with ASD in Mainstream Schools. Procedia - Social and Behavioral Sciences, 46, 4072– 4076. 10.1016/J.SBSPRO.2012.06.200

Gentles, S. J., Ng-Cordell, E. C., Hunsche, M. C., Mcvey, A. J., Dimitra Bednar, E., Chen, Y.-J., Duku, E., Kerns, C. M., Banfield, L., Szatmari, P., & Georgiades, S. (2024). Trajectory research in children with an autism diagnosis: A scoping review. Autism, 28(3), 540–564. 10.1177/13623613231170280

Gotham, K., Pickles, A., & Lord, C. (2009). Standardizing ADOS scores for a measure of severity in autism spectrum disorders. Journal of Autism and Developmental Disorders, 39(5), 693–705. 10.1007/s10803-008-0674-3

Gotham, K., Risi, S., Pickles, A., & Lord, C. (2007). The autism diagnostic observation schedule: Revised algorithms for improved diagnostic validity. Journal of Autism and Developmental Disorders, 37(4), 613–627. 10.1007/s10803-006-0280-1

Harris, S. L., Handleman, J. S., Gordon, R., Kristoff, B., & Fuentes, F. (1991). Changes in cognitive and Language functioning of Preschool children with autism. Journal of Autism and Developmental Disorders, 21(3), 281–290. 10.1007/BF02207325/METRICS

Howlin, P., Moss, P., Savage, S., & Rutter, M. (2013). Social outcomes in mid- to later adulthood among individuals diagnosed with autism and average nonverbal IQ as children. Journal of the American Academy of Child and Adolescent Psychiatry, 52(6), 572–581.e1. 10.1016/j.jaac.2013.02.017

Hus, V., Gotham, K., & Lord, C. (2014). Standardizing ADOS Domain Scores: Separating Severity of Social Affect and Restricted and Repetitive Behaviors. Journal of Autism and Developmental Disorders, 44(10), 2400–2412. 10.1007/s10803-012-1719-1

Hyman, S. L., Levy, S. E., Myers, S. M., Kuo, D. Z., Apkon, C. S., Davidson, L. F., Ellerbeck, K. A., Foster, J. E. A., Noritz, G. H., O’Connor Leppert, M., Saunders, B. S., Stille, C., Yin, L., Brei, T., Davis, B. E., Lipkin, P. H., Norwood, K., Coleman, C., Mann, M., … Paul, L. (2020). Identification, evaluation, and management of children with autism spectrum disorder. Pediatrics, 145(1). 10.1542/PEDS.2019-3447/36917

Ilan, M., Faroy, M., Zachor, D., Manelis, L., Waissengreen, D., Michaelovski, A., Avni, I., Menashe, I., Koller, J., Dinstein, I., & Meiri, G. (2023). Children with autism exhibit similar longitudinal changes in core symptoms when placed in special or mainstream education settings. Autism, 27(6), 1628–1640. 10.1177/13623613221142394

Ilan, M., Meiri, G., Manelis-Baram, L., Faroy, M., Michaelovski, A., Flusser, H., Binoun-Chaki, H., Segev-Cojocaru, R., Dotan, O., Schtaierman, H., Menashe, I., & Dinstein, I. (2021a). Young Autism Spectrum Disorder Children in Special and Mainstream Education Settings Have Similar Behavioral Characteristics. Autism Research, 14(4), 699–708. 10.1002/aur.2400

Ilan, M., Meiri, G., Manelis-Baram, L., Faroy, M., Michaelovski, A., Flusser, H., Binoun-Chaki, H., Segev-Cojocaru, R., Dotan, O., Schtaierman, H., Menashe, I., & Dinstein, I. (2021b). Young Autism Spectrum Disorder Children in Special and Mainstream Education Settings Have Similar Behavioral Characteristics. Autism Research, 14(4), 699–708. 10.1002/aur.2400

Kane, H., & Oakland, T. D. (2015). The differentiation of adaptive behaviours: evidence from high and low performers. Educational Psychology, 35(6), 675–688. 10.1080/01443410.2014.893558

Kim, S. H., Bal, V. H., Benrey, N., Choi, Y. B., Guthrie, W., Colombi, C., & Lord, C. (2018). Variability in Autism Symptom Trajectories Using Repeated Observations From 14 to 36 Months of Age. Journal of the American Academy of Child & Adolescent Psychiatry, 57(11), 837–848.e2. 10.1016/J.JAAC.2018.05.026

Lense, M., Mitchell, E., Hall, C., & Klaiman, C. (2014a). Assessment of Cognitive and Language Abilities in Toddlers with and without Autism Spectrum Disorders: Comparison of the Mullen Scales of Early Learning and the Bayley Scales of Infant and Toddler Development, 3rd Edition.

Lense, M., Mitchell, E., Hall, C., & Klaiman, C. (2014b). Assessment of Cognitive and Language Abilities in Toddlers with and without Autism Spectrum Disorders: Comparison of the Mullen Scales of Early Learning and the Bayley Scales of Infant and Toddler Development, 3rd Edition.

Lindsay, S., Proulx, M., Scott, H., & Thomson, N. (2014). Exploring teachers’ strategies for including children with autism spectrum disorder in mainstream classrooms. Http://Dx.Doi.Org/10.1080/13603116.2012.758320, 18(2), 101–122. 10.1080/13603116.2012.758320

Lord, C., Rutter, M., Di Lavore, P., Risi, S., Gotham, K., & Bishop, S. (2012). Autism and Diagnostic Observation Schedule, Second Edition (ADOS-2) Manual (Part I): Modules 1-4. Western Psychological Services.

Lord, C., Bishop, S. L., & Anderson, D. K. (2015). Developmental trajectories as outcome measures in heterogeneous populations of children with autism spectrum disorders. Journal of Clinical Child & Adolescent Psychology, 44(4), 540–555. 10.1080/15374416.2015.1030752

Luiselli, J., Happé, F., Hurst, H., Freeman, S., Goldstein, G., Mazefsky, C.,, & Newman, D. B. (2013a). Wechsler preschool and primary scale of intelligence. In Ecyclopedia of autism spectrum disorders: Vol. NY: Springer (p. NY: Springer).

Luiselli, J., Happé, F., Hurst, H., Freeman, S., Goldstein, G., Mazefsky, C.,, & Newman, D. B. (2013b). Wechsler preschool and primary scale of intelligence. In F. R. En Volkmar (Ed.), Ecyclopedia of Autism Spectrum Disorders, New York, NY: Springer.

Lynch, S. L., & Irvine, A. N. (2009). International Journal of Inclusive Education Inclusive education and best practice for children with autism spectrum disorder: an integrated approach. 10.1080/13603110802475518

Mandelli, V., Severino, I., Eyler, L., Pierce, K., Courchesne, E., & Lombardo, M. V. (2024). A 3D approach to understanding heterogeneity in early developing autisms. Molecular Autism . 10.1186/s13229-024-00613-5

Ministry of health. (2014). Director-General Circular No. 4/14: Intervention model in special education kindergartens for children with ASD. In journals.sagepub.com (Vol. 15, Issues 1–2). SAGE Publications Inc.

Mouga, S., Bárbara, &, Correia, R., Café, C., Duque, F., & Oliveira, G. (2020). Language Predictors in Autism Spectrum Disorder: Insights from Neurodevelopmental Profile in a Longitudinal Perspective. Journal of Abnormal Child Psychology (2020), 48, 149–161. 10.1007/s10802-019-00578-7

Mullen, E. M. (1995). Mullen scales of early learning. AGS Circle Pines, MN.

Mundy, P., & Newell, L. (2007). Attention, joint attention, and social cognition. Current Directions in Psychological Science, 16(5), 269–274. 10.1111/j.1467-8721.2007.00518.x

Mundy, P., Delgado, C., Block, J., Venezia, M., Hogan, A., & Seibert, J. (2003). Manual for early social communication scales (ESCS). University of Miami. Retrieved from http://www.ucdmc.ucdavis.edu/mindinstitute/ourteam/faculty_staff/escs.pdf

Nahmias, A. S., Kase, C., & Mandell, D. S. (2014). Comparing cognitive outcomes among children with autism spectrum disorders receiving community-based early intervention in one of three placements. Autism, 18(3), 311–320. 10.1177/1362361312467865

Nelson, C. A., Sullivan, E., & Engelstad, A. M. (2024). Annual research review: Early intervention viewed through the lens of developmental neuroscience. Journal of Child Psychology and Psychiatry, 65(4), 435–455. 10.1111/jcpp.13858

Nitzan, T., Bachrach, M., Ilan, M., Faroy, M., Waissengreen, D., Michaelovsky, A., Zagdon, D., Sadaka, Y., Yosef, O. B., Zachor, D., Avni, E., Menashe, I., Meiri, G., Koller, J., & Dinstein, I. (2026). Early joint attention abilities measured by the ADOS-2 predict subsequent expressive language development in minimally verbal autistic children. *JCPP Advances*, e70140. 10.1002/jcv2.70140

Odom, S. L., Hall, L. J., Morin, K. L., Kraemer, B. R., Hume, K. A., McIntyre, N. S., Nowell, S. W., Steinbrenner, J. R., Tomaszewski, B., Sam, A. M., & DaWalt, L. (2021). Educational Interventions for Children and Youth with Autism: A 40-Year Perspective. Journal of Autism and Developmental Disorders, 51(12), 4354–4369. 10.1007/S10803-021-04990-1/METRICS

Oosterling, I., Roos, S., De Bildt, A., Rommelse, N., De Jonge, M., Visser, J., Lappenschaar, M., Swinkels, S., Van Der Gaag, R. J., & Buitelaar, J. (2010). Improved diagnostic validity of the ADOS revised algorithms: A replication study in an independent sample. Journal of Autism and Developmental Disorders, 40(6), 689–703. 10.1007/s10803-009-0915-0

Pope, L., Light, J., & Laubscher, E. (2024). The Effect of Naturalistic Developmental Behavioral Interventions and Aided AAC on the Language Development of Children on the Autism Spectrum with Minimal Speech: A Systematic Review and Meta-analysis. Journal of Autism and Developmental Disorders, 1–22. 10.1007/S10803-024-06382-7/TABLES/4

Rattaz, C., Kerim Munir, ·, Michelon, · Cécile, Picot, M.-C., & Amaria Baghdadli, ·. (2020). School Inclusion in Children and Adolescents with Autism Spectrum Disorders in France: Report from the ELENA French Cohort Study on behalf of ELENA study group. Journal of Autism and Developmental Disorders, 50, 455–466. 10.1007/s10803-019-04273-w

Reed, P., Osborne, L. A., & Waddington, E. M. (2012). BERA A comparative study of the impact of mainstream and special school placement on the behaviour of children with Autism Spectrum Disorders. Educational Research Journal, 38(5), 749–763.

Rojahn, J., Schroeder, S. R., Mayo-Ortega, L., Oyama-Ganiko, R., LeBlanc, J., Marquis, J., & Berke, E. (2013). Validity and Reliability of the Behavior Problems Inventory, the Aberrant Behavior Checklist, and the Repetitive Behavior Scale-Revised among Infants and Toddlers at Risk for Developmental Disabilities: A Multi-Method Assessment Approach. Research in Developmental Disabilities, 34(5), 10.1016/j.ridd.2013.02.024. https://doi.org/10.1016/J.RIDD.2013.02.024

Sandbank, M., Bottema-Beutel, K., Crowley, S., Cassidy, M., Feldman, J. I., Canihuante, M., & Woynaroski, T. (2020). Intervention Effects on Language in Children With Autism: A Project AIM Meta-Analysis. Journal of Speech, Language, and Hearing Research, 63(5), 1537–1560. 10.1044/2020_JSLHR-19-00167

Sandbank, M., Pustejovsky, J. E., Bottema-Beutel, K., Caldwell, N., Feldman, J. I., LaPoint, S. C., & Woynaroski, T. (2024). Determining Associations Between Intervention Amount and Outcomes for Young Autistic Children: A Meta-Analysis. JAMA Pediatrics, 178(8), 763–773. 10.1001/JAMAPEDIATRICS.2024.1832

Sandbank, M., Pustejovsky, J. E., Bottema-Beutel, K., Caldwell, N., Feldman, J. I., LaPoint, S. C., & Woynaroski, T. (2024). Determining associations between intervention amount and outcomes for young autistic children: A meta-analysis. JAMA Pediatrics, 178(8), 763–773. 10.1001/jamapediatrics.2024.1832

Sansour, T., & Bernhard, D. (2018a). ScienceDirect Special needs education and inclusion in Germany and Sweden Enseignement spécialisé et école inclusive en Allemagne et en Suède. European Journal of Disability Research, 12, 127–139. 10.1016/j.alter.2017.12.002

Sansour, T., & Bernhard, D. (2018b). ScienceDirect Special needs education and inclusion in Germany and Sweden Enseignement spécialisé et école inclusive en Allemagne et en Suède. European Journal of Disability Research, 12, 127–139. 10.1016/j.alter.2017.12.002

Stahmer, A. C., & Ingersoll, B. (2004). Inclusive programming for toddlers with autism spectrum disorders: Outcomes from the children’s toddler school. Journal of Positive Behavior Interventions, 6(2), 67–82. 10.1177/10983007040060020201

Sullivan, K., Stone, W. L., & Dawson, G. (2014). Potential neural mechanisms underlying the effectiveness of early intervention for children with autism spectrum disorder. Research in Developmental Disabilities, 35(11), 2921–2932. 10.1016/j.ridd.2014.07.027

Talbott, M. R., Estes, A., Zierhut, C., Dawson, G., & Rogers, S. J. (2016). Early Start Denver Model. 113–149. 10.1007/978-3-319-30925-5_5

Towle, P. O., Vacanti-Shova, K., Higgins-D’Alessandro, A., Ausikaitis, A., & Reynolds, C. (2018). A Longitudinal Study of Children Diagnosed with Autism Spectrum Disorder Before Age Three: School Services at Three Points Time for Three Levels of Outcome Disability. Journal of Autism and Developmental Disorders, 48(11), 3747–3761. 10.1007/S10803-018-3606-X

Trembath, D., & Vivanti, G. (2014). Problematic but predictive: Individual differences in children with autism spectrum disorders. International Journal of Speech-Language Pathology, 16(1), 57–60. 10.3109/17549507.2013.859300

Van Buuren, S., & Groothuis-Oudshoorn, K. (2011). mice: Multivariate Imputation by Chained Equations in R. Journal of Statistical Software, 45(3), 1–67. 10.18637/jss.v045.i03

Van Kessel, R., Walsh, S., Ruigrok, A. N. V., Holt, R., Yliherva, A., Kärnä, E., Moilanen, I., Hjörne, E., Johansson, S. T., Schendel, D., Pedersen, L., Jørgensen, M., Brayne, C., Baron-Cohen, S., & Roman-Urrestarazu, A. (2019). Autism and the right to education in the EU: Policy mapping and scoping review of Nordic countries Denmark, Finland, and Sweden. Molecular Autism, 10(1). 10.1186/s13229-019-0290-4

Viezel, K., Zibulsky, J., Dumont, R., & Willis, J. O. (2014a). Bayley scales of infant and toddler development, third edition. In In C. R. Reynolds, K. J. Vannest, & E. Fletcher-Janzen (Eds.), Encyclopedia of special education: A reference for the education of children, adolescents, and adults disabilities and other exceptional individuals (4th ed.).: Vol. Hoboken (p. NJ: John Wiley & Sons, Inc).

Viezel, K., Zibulsky, J., Dumont, R., & Willis, J. O. (2014b). Bayley scales of infant and toddler development, third edition. *In* *C. R.* *Reynolds*, *K. J.* *Vannest**, &* *E.* *Fletcher-Janzen* *(Eds.),* Encyclopedia of Special Education: A Reference for the Education of Children, Adolescents, and Adults Disabilities and Other Exceptional Individuals (4th Ed.)., Hoboken, NJ: John Wiley & Sons, Inc.

Visser, J. C., Rommelse, N. N. J., Lappenschaar, M., Servatius-Oosterling, I. J., Greven, C. U., & Buitelaar, J. K. (2017). Variation in the Early Trajectories of Autism Symptoms Is Related to the Development of Language, Cognition, and Behavior Problems. Journal of the American Academy of Child and Adolescent Psychiatry, 56(8), 659–668. 10.1016/j.jaac.2017.05.022

Vivanti, G., Dissanayake, C., Duncan, E., Feary, J., Capes, K., Upson, S., Bent, C. A., Rogers, S. J., Hudry, K., Jones, C., Bajwa, H., Marshall, A., Maya, J., Pye, K., Reynolds, J., Rodset, D., & Toscano, G. (2019a). Outcomes of children receiving Group-Early Start Denver Model in an inclusive versus autism-specific setting: A pilot randomized controlled trial. Autism, 23(5), 1165–1175. 10.1177/1362361318801341/ASSET/IMAGES/LARGE/10.1177_1362361318801341-FIG2.JPEG

Vivanti, G., Dissanayake, C., Duncan, E., Feary, J., Capes, K., Upson, S., Bent, C. A., Rogers, S. J., Hudry, K., Jones, C., Bajwa, H., Marshall, A., Maya, J., Pye, K., Reynolds, J., Rodset, D., & Toscano, G. (2019b). Outcomes of children receiving Group-Early Start Denver Model in an inclusive versus autism-specific setting: A pilot randomized controlled trial. Autism, 23(5), 1165–1175. 10.1177/1362361318801341/ASSET/IMAGES/LARGE/10.1177_1362361318801341-FIG2.JPEG

Vivanti, G., Kasari, C., Green, J., Mandell, D., Maye, M., & Hudry, K. (2018). Implementing and Evaluating Early Intervention for Children with Autism: Where Are the Gaps and What Should We Do? International Society for Autism Research, 11, 16–23. 10.1002/aur.1900

Waddington, E. M., & Reed, P. (2017). Comparison of the effects of mainstream and special school on National Curriculum outcomes in children with autism spectrum disorder: an archive-based analysis. Journal of Research in Special Educational Needs, 17(2), 132–142. 10.1111/1471-3802.12368

Wetherby, A. M., Allen, L., Cleary, J., Kublin, K., & Goldstein, H. (2002). Validity and reliability of the communication and symbolic behavior scales developmental profile with very young children. Journal of speech, language, and hearing research, 45(6), 1202–1218.

White, S. W., Scahill, L., Klin, A., Koenig, K., & Volkmar, F. R. (2007). Educational placements and service use patterns of individuals with autism spectrum disorders. Journal of Autism and Developmental Disorders, 37(8), 1403–1412. 10.1007/S10803-006-0281-0

Zachor, D. A., & Ben Itzchak, E. (2010). Treatment approach, autism severity and intervention outcomes in young children. Research in Autism Spectrum Disorders, 4(3), 425–432. 10.1016/J.RASD.2009.10.013

Zachor, Di. A., & Ben-Itzchak, E. (2017). Variables affecting outcome of early intervention in autism spectrum disorder. In Journal of Pediatric Neurology (Vol. 15, Issue 3, pp. 129– 133). Georg Thieme Verlag. 10.1055/s-0037-1601444

Zwaigenbaum, L., Bauman, M. L., Choueiri, R., Kasari, C., Carter, A., Granpeesheh, D., Mailloux, Z., Smith Roley, S., Wagner, S., Fein, D., Pierce, K., Buie, T., Davis, P. A., Newschaffer, C., Robins, D., Wetherby, A., Stone, W. L., Yirmiya, N., Estes, A., … Natowicz, M. R. (2015). Early Intervention for Children With Autism Spectrum Disorder Under 3 Years of Age: Recommendations for Practice and Research:S60-S81. In Pediatrics (Vol. 136). http://publications.aap.org/pediatrics/article-pdf/136/Supplement_1/S60/895851/peds_2014-3667e.pdf

