## Supplementary Material for "Comparing Developmental Outcomes of Autistic Preschoolers Across Special and Mainstream Educational Settings"

**Table S1: Comparison of baseline characteristics between children with and without missing outcome data**

| **Variable** | **Observed (Mean ± SD)** | **Missing (Mean ± SD)** | **Test Statistic** | **p-value** |
|---|---|---|---|---|
| Age at ASD diagnosis | 28.25 (12.85) | 30.57 (6.67) | -1.02 | .314 |
| ADOS age (months) | 39.91 (4.71) | 38.77 (4.76) | 1.14 | .257 |
| Income | 1.56 (1.15) | 1.31 (1.00) | 0.95 | .345 |
| Socioeconomic level | 4.87 (0.90) | 4.66 (1.04) | 1.00 | .319 |
| Maternal education (years) | 13.77 (2.74) | 13.89 (2.47) | -0.22 | .829 |
| Paternal education (years) | 13.63 (2.95) | 13.31 (2.02) | 0.51 | .613 |
| ADOS CSS (baseline) | 6.91 (1.96) | 6.98 (2.23) | -0.17 | .868 |
| ADOS SA CSS (baseline) | 6.44 (2.09) | 6.31 (2.46) | 0.28 | .778 |
| ADOS RRB CSS (baseline) | 7.94 (1.43) | 8.23 (1.59) | -0.92 | .361 |
| Joint attention (baseline) | 7.59 (3.87) | 7.55 (4.09) | 0.04 | .966 |
| Language ability (baseline) | 4.47 (1.91) | 4.98 (1.59) | -1.35 | .184 |
| Cognitive score (baseline) | 70.87 (24.47) | 69.35 (21.01) | 0.28 | .778 |
| ABAS GAC | 68.39 (18.65) | 67.93 (17.22) | 0.11 | .917 |

| Variable | Observed (Mean ± SD) | Missing (Mean ± SD) | Test Statistic | p-value |
|---|---|---|---|---|
| ABAS Conceptual | 71.79 (17.34) | 68.62 (16.15) | 0.78 | .440 |
| ABAS Social | 69.89 (16.73) | 69.44 (16.09) | 0.11 | .911 |
| ABAS Practical | 68.57 (17.55) | 68.51 (17.35) | 0.01 | .989 |
| ABC Irritability | 13.29 (10.46) | 12.93 (11.25) | 0.13 | .896 |
| ABC Social Withdrawal | 9.12 (8.44) | 11.43 (9.96) | -1.01 | .316 |
| ABC Stereotypy | 4.72 (4.66) | 5.14 (5.44) | -0.34 | .737 |
| ABC Hyperactivity | 16.32 (10.31) | 18.20 (12.15) | -0.67 | .506 |
| ABC Inappropriate Speech | 2.92 (2.84) | 3.43 (3.35) | -0.66 | .511 |

**Categorical variables**

| Variable | Test | p-value |
|---|---|---|
| Sex | $\chi^2 = 0.00$ | 1.00 |
| Educational setting | $\chi^2 = 1.12$ | .571 |
